# Transmission is a key driver of extensively drug-resistant tuberculosis

**DOI:** 10.1101/2024.06.28.24309543

**Authors:** Galo A. Goig, Chloé Loiseau, Nino Maghradze, Kakha Mchedlishvili, Teona Avaliani, Daniela Brites, Sonia Borrell, Rusudan Aspindzelashvili, Zaza Avaliani, Maia Kipiani, Nestani Tukvadze, Levan Jugheli, Sebastien Gagneux

**Affiliations:** Swiss Tropical and Public Health Institute, Allschwil, Switzerland; University of Basel, Basel, Switzerland; National Center for Tuberculosis and Lung Diseases (NCTLD), Tbilisi, Georgia; European University, Tbilisi, Georgia; David Tvildiani Medical University (DTMU), Tbilisi, Georgia; The University of Georgia, Tbilisi, Georgia

## Abstract

Multidrug-resistant tuberculosis (MDR-TB) and extensively drug-resistant (XDR) TB are threatening global TB control. The World Health Organization has recently endorsed new regimens for the treatment of MDR-TB that rely on the new and repurposed drugs bedaquiline, pretomanid and linezolid with or without moxifloxacin (BPaL(M)). As BPaL(M) is being rolled-out, resistance to these new drugs is already emerging, leading to acquired XDR-TB. Importantly, instances of transmitted XDR-TB have been reported. The spread of highly drug-resistant *M. tuberculosis* (MTB) strains pose at risk novel TB treatments that took decades to develop. In this study, we analyzed 6,926 MTB genomes from a 13-year nationwide study in Georgia, a known geographical hotspot of MDR-TB, together with more than 80,000 MTB genomes from public sources to estimate the relative contribution of transmission to the burden of XDR-TB. We show that XDR-TB is already geographically widespread, occurring in at least 27 countries across four continents. Moreover, we estimated that a quarter of the XDR-TB cases identified are likely the consequence of transmission. Our findings call for urgent improvements in the global diagnostic capacity, infection control, and surveillance of XDR-TB.

## Introduction

Multidrug-resistant tuberculosis (MDR-TB) and extensively resistant tuberculosis (XDR-TB) are major public health concerns^1^. While drug resistance initially emerges because of multiple factors linked to the quality of TB control and patient adherence, subsequent patient-to-patient transmission (i.e. primary resistance) can contribute to the onward spread of drug-resistant strains of the *Mycobacterium tuberculosis* Complex (MTBC). Indeed, transmission is known to be one of the main drivers of many local epidemics of MDR-TB^2–5^. After more than 40 years without any new TB drug, the World Health Organization (WHO) recently endorsed novel regimens for the treatment of MDR-TB that rely on the new drugs bedaquiline, pretomanid, and the repurposed drug linezolid with or without moxifloxacin (BPaL(M))^6^. As these new regimens are being rolled out, resistance to the new drugs is increasingly being reported^7^. Given the recent introduction of these drugs, most of these cases of XDR-TB are likely due to acquired resistance. Yet, the proportion of XDR-TB transmission has not been determined. Because XDR strains carry many drug resistance-conferring mutations, their reproductive fitness can be reduced, which might also affect their transmissibility^8,9^. However, some MTBC outbreak strains can be both highly drug-resistant and transmissible^8,10^, and a few studies have already reported instances of transmission of XDR-TB^11–13^.

Here, we evaluated the relative contribution of transmission to the burden of XDR-TB by analyzing close to 89,000 MTBC genomes from global sources, and identifying XDR strains based on the catalog of drug resistance-conferring mutations recently released by the WHO^14^. We then identified genomic clusters among these strains, likely indicating transmission of XDR-TB. Our results show that among a total of 514 XDR-TB patient isolates identified across 27 countries, one quarter occurred in genomic clusters with identical mutational profiles. We conclude that XDR-TB is geographically widespread, and that transmission is already an important driver of XDR-TB in different settings.

Our findings call for urgent improvements in the global diagnostic capacity, infection control, and surveillance of XDR-TB, without which, history will repeat itself, jeopardizing the long-awaited treatments for MDR-TB.

## Methods

### Dataset description

Between January 2011 and November 2023, we collected all available MTBC strains from bacteriologically confirmed MDR-TB cases in Georgia. Phenotypic drug resistance was determined at the National Reference Laboratory (NRL) of the National Center for Tuberculosis and Lung Diseases (NCTLD) in Tbilisi, Georgia, using the BD *BACTEC*™ *MGIT*™ 960 system. The process of DNA extraction and whole-genome sequencing has been detailed previously^2,7,8^. Briefly, positive MGIT cultures were subcultured in 7H10 plates. DNA was extracted from three loops of bacterial cells using a phenol-chloroform extraction and subsequently sent for whole-genome sequencing at the genomic core facility of the University of Basel and the Department of Biosystems Science and Engineering at ETHZ in Basel, Switzerland. In total, we compiled 6,926 MTBC genomes from Georgia. Additionally, we screened 81,576 publicly available MTBC genomes. The samples analyzed in this study included previously sequenced genomes as well as new genomes sequenced during this study. Supplementary File 1 contains the accession numbers with additional information for all XDR strains analyzed.

### Ethical approval for samples isolated in Georgia

The institutional Review Board of the NCTLD in Tbilisi, Georgia and the Ethics Commission of North- and Central Switzerland granted ethical approval for this study. The ethics committees waived the need for individual patient consent since only limited and anonymized clinical data were used.

### Bioinformatic analysis: variant calling

All genomes were analyzed following the same in-house analysis pipeline. FASTQ files were processed with Trimmomatic^17^ v0.39 to remove sequencing adapters, trim low quality reads and keep reads longer than 20bp. For paired-end data, reads were merged using SeqPrep^18^ v1.3.1 with an overlap size of 15bp. The resulting reads were mapped to the reconstructed chromosome of the MTBC ancestor^19^ using BWA mem^20^ v0.7.17. Duplicates were identified and removed with Picard^21^ v2.26.2. Sequencing reads were taxonomically classified using Kraken^22^, and non-MTB mappings were discarded as described previously^23^. Local realigments around INDELs was performed using GATK^24^ v4.2.4.1 and variants were called using the microbial mode of GATK Mutect2. Additionally we excluded from analysis supplementary and secondary alignments^25^, and genomic positions in repetitive regions such as PE, PPE, and PGRS genes or phages^26^. Samples with an average sequencing depth lower than 20X or with more than 1% of contaminating reads from non-tuberculous mycobacteria were excluded from downstream analysis.

### Bioinformatic analysis: drug resistance prediction and transmission analysis

To identify drug-resistance (DR) conferring mutations, we compared all mutations detected in each genome with at least 10% allele frequency with the second edition of the catalog released by the WHO^14^. We only considered DR-conferring variants with a final confidence grading of “Assoc w Resistance” or “Assoc w Resistance – Interim”. Mutations in *Rv0678* were considered to confer resistance to bedaquiline only in the absence of loss of function mutations in *mmpL5* as detailed in the WHO catalog^14^. We considered isolates to be XDR if we identified mutations conferring resistance to rifampicin, isoniazid, fluoroquinolones, and at least one of the following: bedaquiline, delamanid, or linezolid. Of note, the WHO catalog does not yet contain any mutations associated with resistance to pretomanid. However, delamanid and pretomanid are both nitroimidazoles, and mutations in genes conferring resistance to delamanid (*fbiA, fbiB, fbiC, ddn* and *fgd1*) can also confer resistance to pretomanid^27^.

For the transmission analysis, we excluded genomes that showed evidence of mixed infection involving different MTBC sublineages. Lineages were identified based on SNPs as described in Coll *et al*.^*28*^. From single-strain infections, we built a multiple alignment of all non-redundant single-nucleotide polymorphisms (SNPs). In this alignment, we only considered SNPs with a minimum intra-sample allele frequency of 90%. Genomic transmission clusters were defined based on a maximum genetic distance of 12 SNPs^29^.

To discriminate between cases in which XDR-TB evolved after transmission of an MDR strain from those in which XDR-TB was likely transmitted from patient to patient, we analyzed the DR-conferring mutations of each sample within each genomic cluster. If more than one DR-conferring mutation was found for a given drug, we only considered the two most prevalent alleles. Genomes within a genomic cluster were only considered to reflect primary transmission of XDR-TB if they shared the same DR-conferring mutations (Supplementary File 1).

## Results and discussion

### Origin of the XDR isolates analyzed and distribution of MTBC lineages

Our screen of a total of 88,502 MTBC genomes, including 6,926 from Georgia and 81,576 from global sources, revealed a total of 514 XDR MTBC genomes from 27 different countries in Africa, Asia, Europe and the Americas (Table 1). For six XDR genomes, the country of isolation could not be determined. Isolates from India (n=179; 35%), South Africa (n=107; 21%), and Georgia (n=60, 11%) contributed to two thirds of all 514 XDR isolates analyzed. Most of the XDR isolates analyzed belonged to MTBC lineage 2 (n=331, 69%), followed by lineage 4 (n=118, 25%), lineage 3 (n=26, 5%) and lineage 1 (n=3, 1%). Among the lineage 2 strains, the majority (n=267; 81%) were “modern Beijing” (L2.2.1) and accounted for 56% of the total dataset. Lineage 4 was the majority lineage only in Mozambique, linked to an outbreak strain from lineage 4.4.1.1 carrying an “escape” *rpoB* variant as described in Barilar *et al*.^13^. XDR lineage 3 were only identified in Bangladesh, India and Pakistan, in line with the global distribution of the main MTBC lineages^31^. Some strains previously linked to large outbreaks of MDR-TB were particularly prominent in certain countries. In particular, L2.2.1 “Central Asia” and L2.2.1 “W148” accounted for most XDR strains in Belarus (n=25; 81%), Georgia (n=42; 70%) and Kazakhstan (n=19; 100%). These outbreak strains are the main drivers of the MDR/XDR epidemic in Eastern Europe and Central Asia, and have been described as carrying a “genetic arsenal” that render them highly drug-resistant and transmissible^8,15,16^.

**Table 1:** Distribution of MTBC lineages according to the country of isolation.

| Country | Lineage 1 | Lineage 2 | Lineage 3 | Lineage 4 | Mixed infection | Total |
| --- | --- | --- | --- | --- | --- | --- |
| Bangladesh | 0 | 2 | 2 | 2 | 1 | 7 |
| Belarus | 0 | 24 | 0 | 6 | 1 | 31 |
| Belgium | 0 | 1 | 0 | 0 | 0 | 1 |
| Brazil | 0 | 0 | 0 | 3 | 0 | 3 |
| China | 0 | 18 | 0 | 0 | 0 | 18 |
| Georgia | 0 | 48 | 0 | 10 | 2 | 60 |
| Germany | 0 | 4 | 0 | 0 | 0 | 4 |
| India | 1 | 114 | 17 | 18 | 29 | 179 |
| Iran | 0 | 0 | 0 | 0 | 1 | 1 |
| Italy | 0 | 0 | 0 | 4 | 0 | 4 |
| Japan | 0 | 1 | 0 | 0 | 0 | 1 |
| Kazakhstan | 0 | 19 | 0 | 0 | 0 | 19 |
| Mexico | 0 | 0 | 0 | 1 | 0 | 1 |
| Moldova | 0 | 3 | 0 | 3 | 0 | 6 |
| Mozambique | 0 | 4 | 0 | 24 | 0 | 28 |
| Myanmar | 0 | 1 | 0 | 0 | 0 | 1 |
| Netherlands | 0 | 0 | 0 | 1 | 0 | 1 |
| Pakistan | 0 | 4 | 7 | 1 | 0 | 12 |
| Peru | 0 | 1 | 0 | 8 | 0 | 9 |
| Portugal | 0 | 0 | 0 | 2 | 0 | 2 |
| Romania | 0 | 0 | 0 | 1 | 0 | 1 |
| Russia | 0 | 5 | 0 | 0 | 0 | 5 |
| Rwanda | 0 | 0 | 0 | 1 | 0 | 1 |
| South Africa | 1 | 71 | 0 | 33 | 2 | 107 |
| South Korea | 0 | 3 | 0 | 0 | 0 | 3 |
| Thailand | 1 | 1 | 0 | 0 | 0 | 2 |
| Ukraine | 0 | 1 | 0 | 0 | 0 | 1 |
| Unknown | 0 | 6 | 0 | 0 | 0 | 6 |
| All dataset | 3 | 331 | 26 | 118 | 36 | 514 |

### Distribution of drug resistance-conferring mutations and totally drug-resistant MTBC strains

By definition, all XDR isolates analyzed were resistant to rifampicin, isoniazid and fluoroquinolones. The distribution of mutations conferring resistance to these drugs followed previous observations across settings. KatG Ser315Thr (n=485; 66%) and Rpob Ser450Leu (n=360; 65%) were the most common isoniazid resistance- and rifampicin resistance-conferring mutations, respectively (Supplementary File 1). Note that the number of drug resistance-conferring mutations analyzed does not necessarily coincide with the number of isolates, given that a particular strain can harbor more than one mutation conferring resistance to a given drug (e.g isoniazid resistance-conferring mutations in both *katG* and *inhA)*. Additionally, 189 (37%) strains carried putative compensatory mutations of rifampicin resistance in RpoA/B/C^9^. The most common mutations conferring resistance to fluoroquionolones were GyrA Asp94Gly (n=244; 41%), Ala90Val (n=112; 19%), Asp94Asn (n=55; 9%) and Asp94Ala (n=51; 9%). The isolates analyzed in this study were also resistant to at least bedaquiline, delamanid or linezolid, alone or in combination, and hence considered XDR. Out of the 514 XDR strains analyzed, 362 (70%) had mutations conferring resistance to bedaquiline, 93 (18%) had mutations conferring resistance to delamanid/pretomanid, and 186 (36%) had mutations conferring resistance to linezolid according to the WHO catalog^14^. Most bedaquiline resistance-conferring mutations occurred in *Rv0678* (n=427; 91%), but mutations were diverse, in most cases leading to frameshifts or premature stop codons (Supplementary File 1). 7% of mutations occurred in *pepQ* and only 2% in *atpE*. The most common delamanid/pretomanid resistance-conferring mutations were in the *fbiA/B/C* genes (n=76; 60%), followed by mutations in *ddn* (n=31; 24%). In the case of linezolid, the most common mutation was Cys154Arg in RplC (n=145; 72%). Because rates of baseline linezolid resistance in untreated patients have been shown to be low^32^, the reason why this mutation is so frequent might be that it has higher chances of evolving *de novo* than other mutations conferring resistance to linezolid^33^. This observation motivated one of the exclusion criteria in the sensitivity analysis described below.

Importantly, we identified nine strains showing combined resistance to bedaquiline, delamanid/pretomanid and linezolid. These strains can be considered “totally” drug-resistant (TDR), and were isolated in Belarus (n=2), Georgia (n=3), India (n=3) and South Africa (n=1). Five of these strains were found in genomic clusters. However, inspection of their detailed mutational profiles suggested that none of these cases involved direct transmission of TDR, but that TDR evolved in all cases after transmission of an XDR strain (Supplementary File 1). Five out of nine TDR strains (56%) carried compensatory mutations of rifampicin resistance. All TDR strains with compensatory mutations were within genomic clusters, with the exception of one mixed infection that was excluded from transmission analysis.

### Distribution of cluster sizes and detailed description of genomic clusters from Georgia

In Georgia, we identified 60 XDR-TB cases, two of which showed evidence of mixed infection. Out of the 58 cases with single-strain infections, 20 cases were in one of six genomic clusters. The median size of these clusters was 3.5 (interquartile range = 2.25 – 4). Analysis of the mutational profile of these clusters suggested that four clusters involving 16/58 patients (28%) were linked to direct transmission of XDR-TB (Supplementary File 1).

One cluster of four patients was defined by the bedaquiline resistance-conferring mutation AtpE Ile66Met. The first diagnosed patient in 2017 was a new TB case, and had prior contact with another drug-resistant case, already indicating primary resistance. This patient was treated with a regimen containing bedaquiline, linezolid and delamanid, which most likely was not efficacious. Importantly, among the three likely secondary cases, one case acquired additional mutations conferring resistance to linezolid (RplC Cys154Arg) and delamanid/pretomanid (loss of function mutation in *fbiC*), therefore qualifying as TDR.

Two other clusters were defined by frameshift mutations in *Rv0678* conferring resistance to bedaquiline. One cluster involved three patients diagnosed in 2022, including one patient with a previous history of incarceration. One patient withdrew from a 9-month treatment, which included bedaquiline and linezolid. The other two received bedaquiline, linezolid and delamanid for 18 months and were cured. The other cluster involved five patients, with the first case diagnosed in 2016 and the remaining in 2022. The case diagnosed in 2016 had a previous episode of TB ending in treatment failure and was treated with a regimen containing bedaquiline, linezolid and clofazimine.

The remaining cluster of four patients was defined by a delamanid resistance-conferring mutation leading to a premature stop codon in the 88^th^ residue of Ddn (Trp88*). The first case was diagnosed in 2012 and had prior contact with a DR-TB case. Two other cases were diagnosed in 2014 and the last one in 2021. As observed in a large cluster in Mozambique, we also found evidence that delamanid resistance could have evolved before fluoroquinolone resistance, given that all four strains carried different *gyrA* mutations. However, the mutation Trp88* in *ddn* is graded as “Assoc w Resistance interim” only under relaxed thresholds in the WHO catalog^14^. Because the case diagnosed in 2012 already had this mutation without a previous history of treatment with delamanid or pretomanid, an alternative explanation is that the *ddn* Trp88* mutation does not confer resistance to these drugs and that this is actually an MDR-TB cluster.

Across samples from global sources, excluding samples from Georgia, 141 out of 420 single-strain XDR-TB infections (34%) were within one of 50 genomic clusters. In most cases, genomic clusters involved two patients (median=2, interquartile range=2-2). After analyzing the detailed drug resistance mutational profile of each cluster, we determined that 41 involving 117 cases (28%) were linked to direct transmission of XDR-TB. The largest cluster showing evidence of direct transmission of XDR-TB involved ten patients from Mozambique. The transmitted strain responsible for this cluster is a genotype carrying the uncommon rifampicin-resistant variant RpoB Ile491Phe that is not detected by GenXpert^13^. This cluster was described previously by Barilar *et al*.^*13*^ as a cluster involving thirteen patients. In our analysis, this cluster showed two separate genomic clusters of ten and three patients, respectively. This is probably due to differences in the bioinformatics analysis pipeline, given that the minimum distance between samples from both clusters was of 13 SNPs, only one SNP above the clustering threshold. Interestingly, while all the strains in this cluster shared the same bedaquiline resistance-conferring mutation (*Rv0678* Met146Thr), they carried different mutations in *gyrA* conferring resistance to fluoroquinolones. Because this mutation is only observed in other two samples across the entire dataset, this observation suggests that bedaquiline resistance evolved before fluoroquinolone resistance, maybe as a consequence of clofazimine cross-resistance. An alternative, yet very unlikely explanation, is that the same bedaquiline resistance mutation evolved *de novo* in samples from the same cluster.

Interestingly, we only observed one genomic cluster of three MTBC lineage 3 XDR strains. The rate of clustering was significantly lower for lineage 3 strains (3/26; 12%) as compared to lineage 2 strains (114/331; 34%; Fisher’s exact test p-value = 0.016) and lineage 4 strains (44/118; 37%; Fisher’s exact test p-value = 0.011). Because the dataset analyzed does not originate from a population-based study, and because only 26 XDR strains belonged to MTBC lineage 3, the difference in clustering rates should be interpreted with caution. However, already the fact that in a dataset of XDR strains only few belong to MTBC lineages 1 and 3, particularly given that in some countries these are the predominant circulating lineages^31^, suggests that transmission of XDR-TB may be influenced by the bacterial genetic background. Additionally, these observations are in line with previous evidence, that has shown local epidemics of drug-resistant TB to be caused mainly by MBTC lineage 2 and 4^2-5,8,11-13,15,16^.

### Sensitivity analysis of transmitted drug resistance

During our analysis, we noted that some publicly available MTBC genomes could have originated from the same patient, which would inflate the inferred number of cases with transmitted XDR-TB. Hence, we excluded these instances based on the metadata provided for each study, the descriptions in the original manuscripts, or obtaining direct confirmation from the respective authors when possible. However, the dataset analyzed may still include strains that could have originated from the same patient due to either the absence of patient identifiers or possible overlaps between studies. This was particularly true for genomes derived from the CRyPTIC project (214 isolates, 42% of the dataset), given that different laboratories across the world have contributed to this project, and no patient-related data were available^34^. Another potential bias in our study was defining direct transmission of drug resistance based on resistance-conferring mutations that are likely to emerge frequently in parallel (i.e. showing high homoplasy). As explained above, we observed that the mutation Cys154Arg in RplC is the most common mutation conferring resistance to linezolid. If transmission of resistance is inferred solely based on this mutation, independent evolution of this particular mutation in two clustered cases may be misinterpreted as transmitted resistance. Therefore, we conducted a more stringent sensitivity analysis by excluding any clustered isolates: i) derived from the CRyPTIC consortium, ii) showing a possible overlap between studies, iii) showing the possibility of being isolated from the same patient, and iv) where transmitted XDR-TB was based solely on RplC Cys154Arg (Supplementary File, column “sensitivity analysis”). Following these criteria, only 62 clustered isolates could be analyzed. Out of these, 53 isolates (85%) in one of 15 genomic clusters likely represented direct transmission of XDR-TB. Therefore, even based on this highly conservative analysis, at least 14% of XDR-TB isolates were likely the result of direct transmission of XDR-TB involving seven countries (Bangladesh, China, Georgia, India, Moldova, Mozambique and Russia). We note that the results of this sensitivity analysis represent the lower bound of our estimations, but it is more likely that they represent a gross underestimation, given the stringency of the criteria used. A clear indicator is the fact that our estimation of the contribution of transmission to the burden of XDR-TB in the “global dataset” (28%) was the same as the estimation in the dataset from Georgia (28%), that stems from a well-sampled, population-based study of MDR-TB spanning 13 years. We also note that if the mutation in Ddn Trp88* does not confer delamanid resistance, then the cluster of four patients in Georgia carrying this mutation would not qualify as XDR-TB, and hence the contribution of transmission to the burden of XDR-TB in Georgia would be 20%.

## Data Availability

Supplementary File 1 contains the accession numbers with additional information for all XDR strains analyzed.

## Acknowledgments

This work was supported by the Swiss National Science Foundation (grants 320030-227432, and CRSII5_213514) and the European Research Council (883582-ECOEVODRTB). Calculations were performed at sciCORE (http://scicore.unibas.ch/) scientific computing core facility at the University of Basel and on the Euler cluster at ETH Zürich. Sequencing was carried out at the Genomics Facility Basel of the University of Basel and the Department of Biosystems Science and Engineering at ETHZ in Basel, Switzerland.

